# KAN-EEG: Towards Replacing Backbone-MLP for an Effective Seizure Detection System

**DOI:** 10.1101/2024.06.05.24308471

**Authors:** Luis Fernando Herbozo Contreras, Jiashuo Cui, Leping Yu, Zhaojing Huang, Armin Nikpour, Omid Kavehei

**Author notes:** Luis Fernando Herbozo Contreras and Jiashuo Cui contributed equally to this work (e-mail: {, }).

## Abstract

The landscape of artificial intelligence (AI) research is witnessing a transformative shift with the emergence of the Kolmogorov-Arnold Network (KAN), presenting a novel architectural paradigm aimed to redefine the structural foundations of AI models, which are based on Multilayer Perceptron (MLP). Through rigorous experimentation and meticulous evaluation, we introduce the KAN-EEG model, a tailored design for efficient seizure detection. Our proposed network is tested and successfully generalized on three different datasets, one from the USA, one from Europe, and one from Oceania, recorded with different front-end hardware. All datasets are scalp Electroencephalogram (EEG) in adults and are from patients living with epilepsy. Our empirical findings reveal that while both architectures demonstrate commendable performance in seizure detection, the KAN model exhibits high-level out-of-sample generalization across datasets from diverse geographical regions, underscoring its inherent adaptability and efficacy at the backbone level. Furthermore, we demonstrate the resilience of the KAN architecture to model size reduction and shallow network configurations, highlighting its versatility and efficiency by preventing over-fitting insample datasets. This study advances our understanding of innovative neural network architectures and underscores the pioneering potential of KANs in critical domains such as medical diagnostics.

## I. Introduction

EPILEPSY is a neurological disorder causing recurring seizures, affecting millions globally. In about 30-35% of cases, standard anti-epileptic drugs (AEDs) fail to control abnormal brain activities, resulting in drug-resistant epilepsy. Despite advancements in AED development and testing, improvements in their effectiveness have been limited [1]. Unpredictable and unprovoked seizures significantly impact patients’ quality of life, employment, and overall well-being, posing risks such as falls and sudden unexpected death in epilepsy [2], [3]. An accurate system for detecting and counting seizures can greatly enhance decision-making, treatment planning, and disease management, leading to better patient outcomes. Electroencephalogram (EEG) signals are commonly used by scientists to diagnose neurological diseases such as seizures. It is a technique that records brain electrical activity by placing electrodes on the scalp to capture electrical signals generated by neuronal activity. [4]. These electrical signals reflect the brain’s functional state and information processing, making EEG an important tool for studying brain function [5]. Variations in EEG signals in time and frequency can reveal brain activity patterns associated with sleep, cognition, emotion, and pathological states such as epilepsy [6]. Artificial intelligence (AI) has undeniably made significant strides in healthcare, becoming a new era of innovation and patient care. Its impact on the healthcare industry has been overwhelmingly positive, with numerous benefits spanning various aspects of medical practice, research, and patient outcomes [7]–[9]. Among the various AI techniques, machine learning models, particularly neural networks, have shown great promise in analyzing and interpreting complex medical data.

Multilayer Perceptron (MLP) networks are the backbones of today’s AI architectures [10], [11], and has been extensively used for its effectiveness in detecting and classifying abnormalities from biosignals. The MLP, a feed-forward artificial neural network, consists of multiple layers of neurons that process input data through weighted connections and activation functions. It excels in capturing the relationships within the data, making it well-suited for analyzing the intricate patterns present in medical signals. Despite its advantages, the MLP has limitations, particularly in model interpretability and efficiency. To address these challenges, a new model known as the Kolmogorov-Arnold Network (KAN) has been proposed as a promising alternative to MLPs [12]. Unlike MLPs, which use fixed activation functions on the hidden layers, KANs employ learnable activation functions, replacing linear weights with univariate functions parametrized as splines. This architectural difference allows them to achieve greater accuracy and interoperability.

KANs have demonstrated the ability to outperform MLPs with smaller network sizes, making them more computationally efficient [12]. Despite its advantages, the MLP has limitations, particularly in model interpretability and efficiency. Furthermore, it has demonstrated the ability to outperform MLPs with smaller network sizes, making them more computationally efficient [12]. In the context of EEG signal analysis, KANs offer potential advantages over MLPs. Their learnable activation functions and efficient representation of data structures enable better handling of the complex, high-dimensional data characteristic of EEG signals. This can improve detection accuracy and more insightful interpretations of the underlying neural activity. The subsequent sections will delve deeper into the comparative performance of our proposed architectures and MLPs in detecting epileptic seizures, highlighting the strengths and potential of this critical application. The diagram in Fig. 1 delineates the structural differences between the KAN and the MLP architectures. The KAN structure demonstrates learnable activation functions, which can constitute interpretable systems and face MLP architecture challenges by enhanced capabilities for continual learning and efficiency at shallow, sparse connections.

**Fig. 1.**
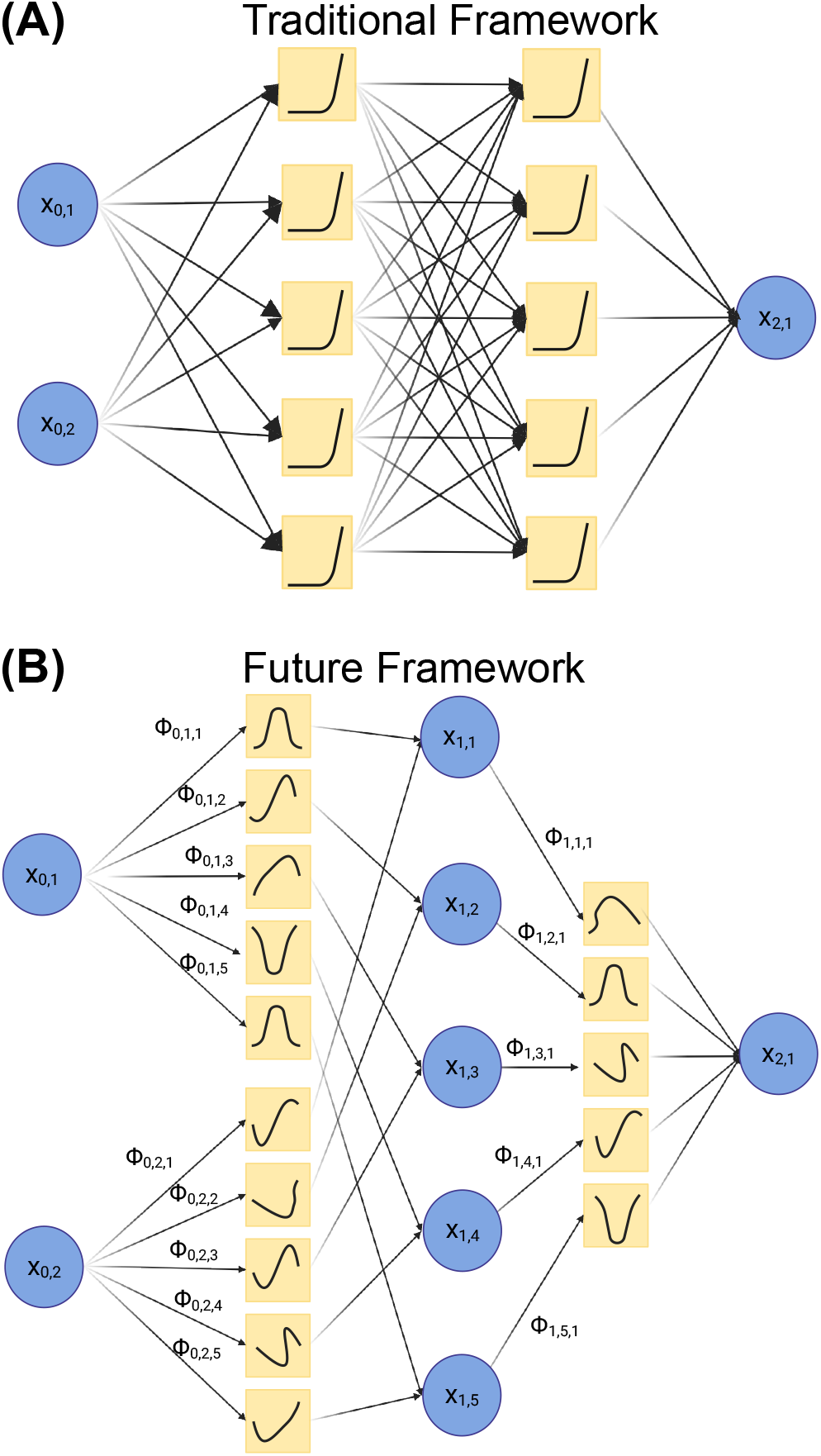
Representation of the past and future in neuro-AI. The multilayer perceptron was built based on non-learnable activation functions and relies on deep, fully connected networks for accurate performance (A). On the contrary, KAN emerges as a potential solution for more explainable, shallow, and efficient architectures at the core level (B).

### A. Background

Epileptic seizure detection has seen substantial progress with the rise of machine learning models, particularly those utilizing EEG signals. EEG tests record the electrical activity produced by neurons, typically noninvasively, by placing multiple electrodes on the scalp [4]. EEGs play a crucial role in detecting epileptic foci and categorizing epilepsy types, such as focal, generalized, and unknown seizures [13]–[15]. These recordings are essential for diagnosing epilepsy, monitoring ongoing conditions, making predictions, and effectively responsive neurostimulation. Recent advancements in deep learning have introduced various architectures to improve EEG-based seizure detection. Convolutional Neural Networks (CNNs) are prominently used, transforming EEG signals into different dimensional forms for detailed analysis [16]–[21]. Recurrent Neural Networks (RNNs), including their advanced versions like Long Short-Term Memory (LSTM) and Gated Recurrent Units (GRU), are adept at capturing temporal dependencies in EEG data, making them suitable for sequential data analysis [22]–[25]. Unsupervised learning methods, such as Deep Belief Networks (DBNs) [26], [27] and Auto-Enconders (AEs) [28]–[30], are employed to extract and reconstruct features from raw signals.

Table I summarizes various traditional models applied to different bio-signal applications, preprocessing methods, datasets, and indicators. In addition, hybrid models that combine CNNs with RNNs or AEs leverage the spatial feature extraction capabilities of CNNs and the temporal modeling strengths of RNNs or the reconstruction abilities of AEs [31]– [36]. For example, Convolutional Long Short-Term Memory (ConvLSTM) networks integrate the spatial feature extraction power of CNNs with the temporal sequence modeling capabilities of LSTMs [31]. This integration has enhanced the model’s performance in detecting seizures.

**TABLE I.** Different types of neural networks applied to EEG signals for epileptic seizure detection.

| Model | Application | Pre-Processing <sup>+</sup> | Dataset <sup>◇</sup> | Metrics <sup>†</sup> |
| --- | --- | --- | --- | --- |
| 3D-CNN + RNN [34] | Seizure detection | STFT, 2D-mapping | CHB-MIT<br>SNUH | ACC: 99.4%<br>ACC: 99.2% |
| ConvLSTM [31] | Seizure detection | ICA, STFT | TUH<br>EPILEPSIAE<br>RPAH | AUC: 84%<br>AUC: 81%<br>AUC: 82% |
| SNN + ConvLSTM [42] | Seizure detection | STFT | FB<br>CHB-MIT<br>EPILEPSIAE | AUC: 92.7%<br>AUC: 89%<br>AUC: 81.1% |
| TSD [37] | Seizure detection | STFT | TUH | AUC: 92.1% |
| ResNet-LSTM [43] | Seizure detection | ICA, STFT | U-Bonn | ACC: 97% |
| GraphS4mer [44] | Seizure detection<br>Sleep staging<br>ECG classification | STFT | TUH<br>DOD-H<br>ICBEB | AUC: 90.6%<br>Macro-F1: 81.8%<br>AUC: 97.7% |
| SVM [45] | Seizure detection | DWT | CHB-MIT | ACC: 98% |

<sup>+</sup> Pre-Processing methods: Short-Time Fourier Transform (STFT), Independent Component Analysis (ICA), Discrete Wavelet Transform (DWT).
<sup>◇</sup> Datasets: Temple University Hospital (TUH), Children’s Hospital Boston (CHB-MIT), Royal Prince Alfred Hospital (RPAH), Seoul National University Hospital (SNUH), University of Bonn (U-Bonn), Freiburg (FB), DREAM Open Dataset-Healthy (DOD-H), International Conference on Biomedical Engineering and Biotechnology (ICBEB).
<sup>†</sup> Metrics: Accuracy (ACC), Area Under the Curve (AUC), Macro-Averaged F1-Score (Macro-F1).

Transformer-based networks, a more recent innovation, incorporate attention mechanisms to better capture complex patterns in EEG data, further improving detection accuracy [37]–[40]. Despite these significant advancements, traditional seizure detection models still encounter challenges in generalization and real-time implementation. Models trained on large datasets often struggle with low Area Under the Receiver Operating Curve (AUROC) and high false positive rates, which limits their clinical applicability. Thus, there is a pressing need for models that can balance sensitivity and specificity and be broadly applicable across diverse patient populations and real-world scenarios [31], [41].

Although the traditional MLP-based model has significantly contributed to the development of EEG-based seizure detection systems, the emergence of KAN and other advanced models provides a promising direction for improving the accuracy, efficiency, and applicability of epilepsy detection technology. The following section will provide a detailed comparison between MLP and KAN, emphasizing the advantages and potential of KAN in EEG signal analysis of epilepsy detection.

### B. Novelty and significance

In this study, our primary aim is to introduce a novel backbone architecture for advancing the field of seizure detection, focusing on addressing challenges related to generalization and real-time implementation. We propose the utilization of KANs as a key component for identifying epileptic seizures from pre-recorded EEG signals. We incorporate the short-time Fourier transform (STFT) technique to process the EEG data effectively. Rather than solely emphasizing the challenges in generalization and implementation, we highlight the innovative potential of efficient-shallow architectures that are adaptable as a foundational framework for future AI models in seizure detection. Through our experimentation with the 3 different continental datasets, we aim to showcase the inherent efficacy of KANs and their capacity to serve as an efficient architecture for developing advanced seizure detection systems.

- **Higher Accuracy**: Preliminary findings suggest that our model demonstrates comparable AUROC values to traditional MLP models, indicating a similar level of performance in seizure detection. This comparative analysis suggests that while KANs do not necessarily outperform MLP models, they offer a comparable level of reliability, thereby contributing to the advancement of seizure detection methodologies.
- **Efficiency**: Our proposed architecture requires a smaller network size. Thereby making them computationally efficient and suitable for real-time applications.
- **Out-of-sample seizure detection**: Generalization beyond the training dataset was assessed by training our model on the USA dataset and evaluating its performance on independent datasets from Europe and Oceania, using the same trained weights. The outcomes indicate encouraging support for this test.

## II. Methods

### A. Datasets

Three datasets were used in this work: the Temple University Hospital (TUH) EEG Corpus, the scalp-EPILEPSIAE dataset, and the Royal Prince Alfred Hospital (RPAH) dataset. Fig. 2 summarizes the TUH dataset, detailing key statistics. This dataset is divided into training and validation sets, offering comprehensive insights into the number of hours utilized for both patients with seizures and non-seizures and the quantity of patients that present seizures and non-seizures. The TUH dataset is the primary training dataset, providing a diverse range of seizure signals for various seizure types. With its large volume of files, the TUH dataset from America (USA) offers extensive data for robust model training.We utilized 400 hours of EEG data for our training process, comprising 120,000 samples with a 12-second window. This dataset included approximately 75% background activity and 25% seizure activity. Notably, this represents a significantly smaller data volume than other models. Despite the reduced dataset size, we successfully validated the KAN-EEG model’s efficiency in seizure detection with reduced training data. The validation was conducted using 192 hours of EEG data, corresponding to 57,306 samples with a 12-second window, and maintained the same 75%-25% ratio of background to seizure information.

**Fig. 2.**
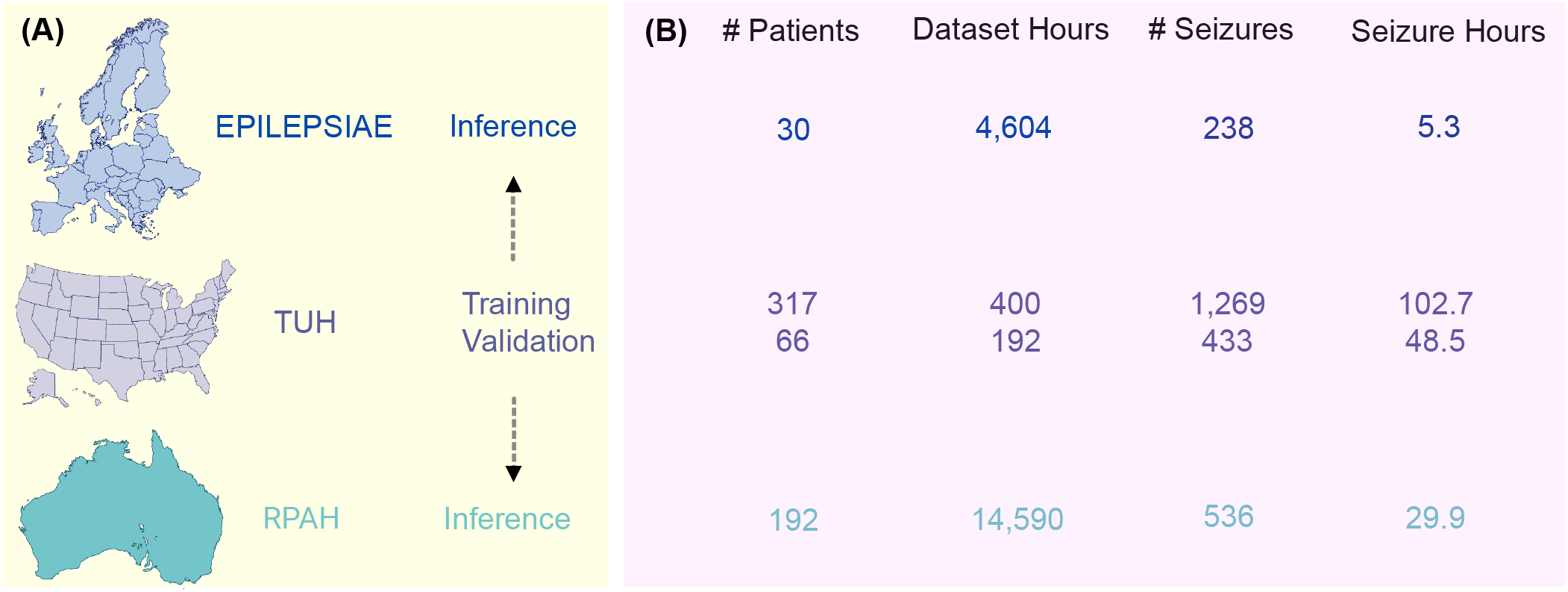
Summary of the datasets being used. The TUH dataset was set as the training and validation dataset and subsequently used for inference in both RPAH and EPILEPSIAE datasets (A). Testing across all datasets incorporates extensive background and seizure session data, facilitating comprehensive analysis of model efficacy in diverse clinical settings (B).

The EPILEPSIAE and RPAH datasets, consisting of adult EEG, are used for inference tests. Both datasets share common characteristics, such as using identical montages and adult patient data, ensuring consistency and comparability in analysis. Integrating these complementary datasets enhances the comprehensiveness and reliability of the AI model for seizure detection. The EPILEPSIAE dataset consists of 30 patients, where 19 and 11 are male and female, respectively. The RPAH dataset from one of Australia’s major hospitals reliably maintained one of the largest datasets from adult epilepsy patients nationwide. This work uses nine years (2011–2019) of data, testing nearly 14,590 hours of EEG data from 192 patients over 1,006 sessions, each averaging around 15 hours of recording. Out of 212 patients, 20 were excluded for reasons including excessive seizures (more than 11 seizures/24 hours), missing electrode data, or seizures confirmed only by video.

The Australian dataset is about 16 times larger than the US training dataset, with longer inter-ictal periods and background data, making evaluating false positives highly robust. The distribution of the RPAH dataset across three domains—seizure type and frequency, age and gender, and seizure occurrence within a 24-hour cycle—highlights essential patterns. Notably, seizure occurrences are derived from intermittent monitoring, providing insights into the likely timing of seizures. Ethical approval was obtained to access this clinical data.

### B. The KAN-EEG Structure

#### 1) Characteristics of KANs

KANs are founded on the Kolmogorov-Arnold representation theorem, which posits that any multivariate continuous function can be broken down into a finite composition of continuous univariate functions and addition operations [12]. This foundational principle allows it to substitute traditional linear weights with spline-parametrized univariate functions. Moreover, it employs adaptive univariate activation functions along the network edges, enhancing flexibility and precision. These functions adjust based on the data, leading to more accurate approximations. Spline functions enable dynamic adaptation to the data, providing refined representations that effectively capture smooth transitions. Additionally, KANs require fewer parameters than MLPs, improving computational efficiency and model interpretability. The learnable functions can also be visualized for better understanding. The operation of a KAN layer can be described by the Equ. 1:

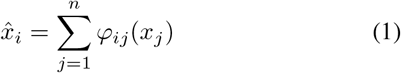

where 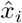 represents the activation value at node *i*, and *φ*_*ij*_ denotes the learnable activation function on the edge connecting node *j* to node *i*.

#### 2) Mathematical Formulation of KANs

KANs utilize the Kolmogorov-Arnold representation theorem to break down a high-dimensional function into a sum of univariate functions. This decomposition is given by the Equ. 2:

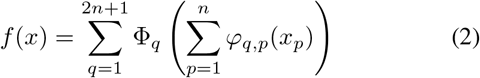

In this formulation, Φ_*q*_ and *φ*_*q,p*_ are univariate functions parameterized as splines, while *x*_*p*_ represents the input features. The inner functions *φ*_*q,p*_(*x*_*p*_) transform the input features into intermediate representations, which are then aggregated and processed by the outer functions Φ_*q*_. This structured approach enables KANs to effectively capture both compositional structures and univariate functions, providing a robust framework for function approximation [12]. An overall representation of the KAN structure is seen in Fig. 1.

### C. Pre-Processing

We utilized two signal processing techniques, Independent Component Analysis (ICA) and Short-Time Fourier Transform (STFT), to address the challenges associated with raw EEG data. Initially, the EEG signals were divided into 12-second segments, and the ICA algorithm was applied to decompose the signals into 19 independent components using Blind Source Separation (BSS). ICA separates EEG signals into statistically independent components, as represented in Equ. (3),

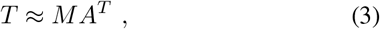

where *T* contains the EEG data, *M* contains the time information, and *A* contains the weights for topographic maps. Pearson correlation was used to identify independent sources strongly associated with eye movement, detected from the ‘FP1’ and ‘FP2’ EEG channels. These sources related to eye movement were removed, resulting in EEG signals free from such artifacts. Subsequently, the Short-Time Fourier Transform (STFT) was applied to the cleaned EEG signals. This involved using a window length of 250 samples (equivalent to 1 second) with a 50% overlap and eliminating the DC component of the transform. As a result, the data dimensions were (*N* × 23 × 125), where *N* represents the number of electrodes, 23 represents the time index, and 125 represents the frequencies. Data preprocessing is performed separately from the KAN model, ensuring the data is adequately prepared before input into the KAN model for further training/inference, as depicted in Fig. 3.

**Fig. 3.**
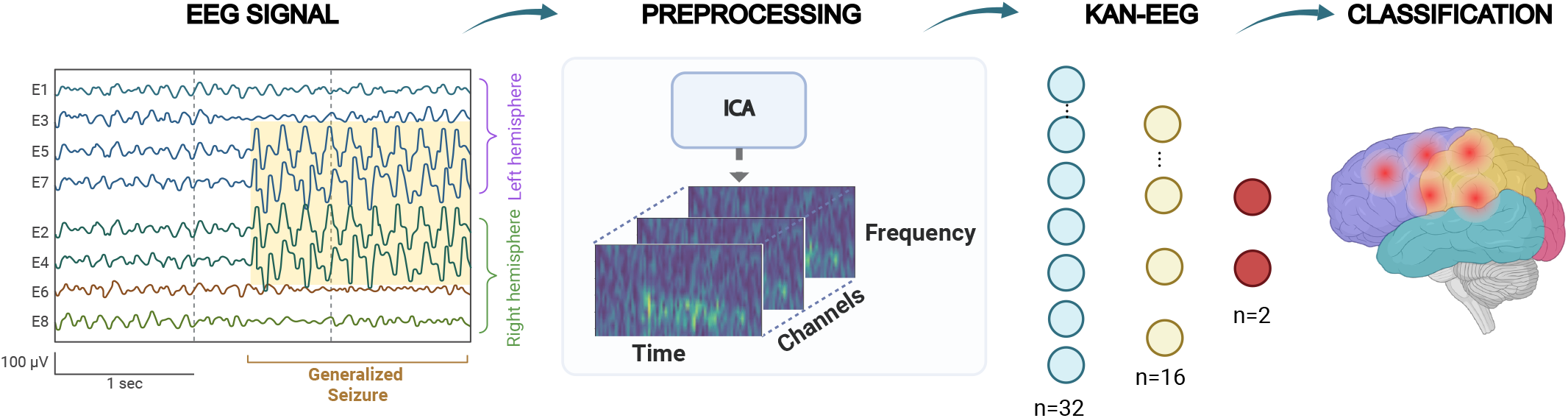
KAN-EEG Seizure System. We analyzed different kinds of seizures utilizing the TUH dataset for training. We then preprocessed our model utilizing Independent Component Analysis (ICA) and Short-Time Fourier Transform (STFT). Subsequently, we incorporate a shallow KAN algorithm that leads to efficient results.

## III. Experiments and Results

### A. Training and Validation in Sample

Our model was trained and validated using the TUH dataset. We achieved an impressive AUROC score of 0.89 in Fig. 4. It’s important to highlight that our model was trained on 400 hours of data, considerably fewer than the 752 hours used for the ConvLSTM model and the 910 hours for the Transformer model. These preliminary results highlight the exceptional performance of our approach, which not only rivals but also exceeds that of contemporary methods by considering a smaller training dataset but a comparable and even greater test dataset. Our model demonstrates superior efficacy in accurately detecting epileptic seizures, underscoring its potential for clinical applications. The structure used to obtain the outlined results is highlighted in Table II, and we use this as our baseline for our next assessment.

**TABLE II.** Model assessment robustness in-sample validation using TUH dataset by decreasing layers and neurons leads to efficiency.

| Model Structure | Layers | Hidden Neurons | Neuron Reduction | Model Size Reduction | AUROC | AUPRC | Precision | Recall | F1-Score |
| --- | --- | --- | --- | --- | --- | --- | --- | --- | --- |
| **I-764-256-O<sup>◊</sup>** (Large) | 2 | 1020 | **Baseline** | **Baseline** | 88.9 | 77.4 | 85.0 | 84.7 | 85.1 |
| **I-32-32-O<sup>\*</sup>** | 2 | 64 | 6.27 % | 4.5 % | 86.9 | 73.5 | 83.2 | 83.0 | 83.3 |
| **I-32-16-O<sup>‡</sup>** (Small) | 2 | 48 | 4.71 % | 4.38 % | 86.7 | 73.1 | 82.9 | 83.1 | 82.8 |
| **I-256-O** | 1 | 256 | 25 % | 29.8 % | 85.3 | 70.1 | 81.3 | 81.2 | 80.9 |

**I**: Input Data. We utilized 23 time and 125 frequency bins, followed by 19 EEG channels.
**O**: Output Neurons. We utilized 2 output neurons for representing states of seizure and non-seizure.
<sup>◊</sup> Out-of-sample generalizations are shown in Tables III and IV.
<sup>\*</sup> Out-of-sample generalizations are shown in Tables III and IV.
<sup>‡</sup> Out-of-sample generalizations are shown in Tables III and IV.

**Fig. 4.**
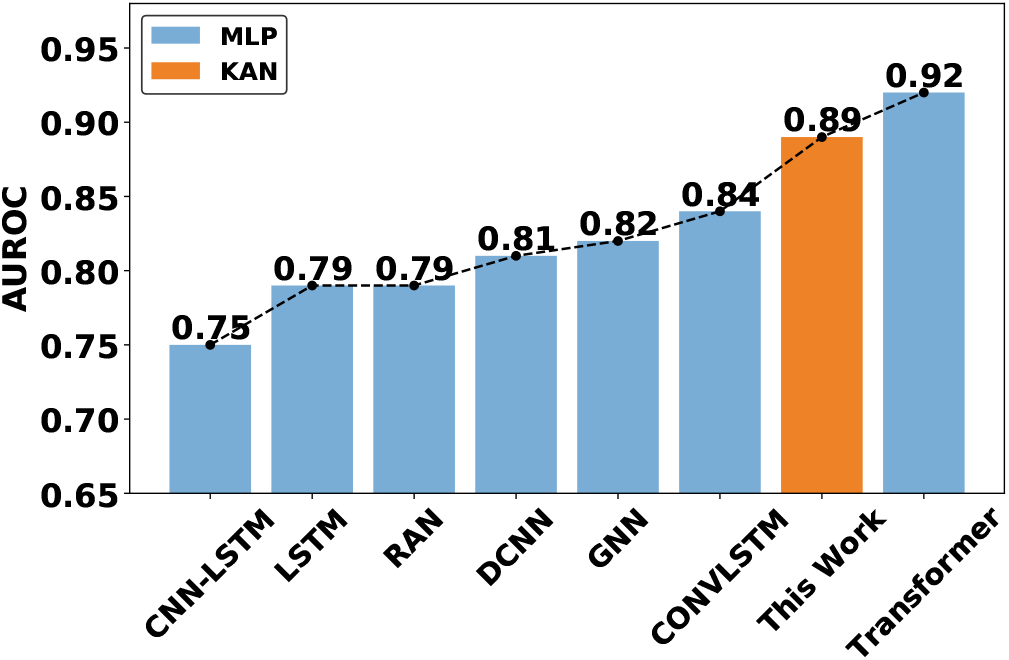
Comparison of KAN-EEG (^***°***^Architecture **I-764-256-O** [see Table II]) training on TUH dataset with other MLP based models [31], [37], [46]–[48]. Our model was trained on 400 hours of data, which is significantly less than the amount of data used to train other models, such as ConvLSTM and Transformer, with 752 hours and 910 hours, respectively.

The training processing was performed for 100 epochs, where in Fig. 5 can be seen the trend across different metrics. Among these, the discernible reduction in the training loss is evident, gradually converging towards optimal thresholds. Noteworthy is the observation that metrics such as Recall, Precision, and AUROC manifest a discernible tendency to reach the convergence phase approximately around the 20^th^ epoch. Training losses have an average convergence between 0.2 and 0.3, given the nature of a small architecture.

**Fig. 5.**
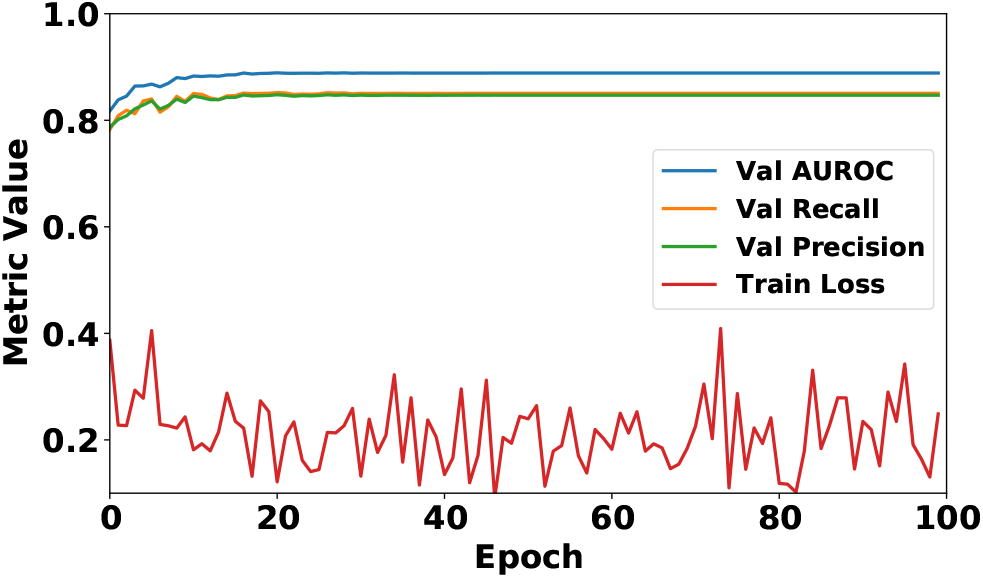
Validation process utilizing the KAN-EEG on TUH dataset.

### B. Robustness

The findings presented in Table II offer compelling insights into the robustness of the KAN-EEG architecture in seizure detection. By systematically reducing the complexity of the model through decreases in both layers and hidden neurons, we sought to evaluate the architecture’s adaptability and efficiency under varying structural configurations. Our analysis reveals that despite the reduction in model complexity, the KAN architecture consistently maintains a high level performance across key evaluation metrics, as reflected by the consistently high AUROC values across all model variants. This indicates the model’s ability to effectively differentiate between seizure and non-seizure states, even when the architectural complexity is significantly reduced.

Moreover, while marginal decreases are observed in metrics such as precision and recall with model simplification, the overall performance, as quantified by the F1-Score, remains robust. This suggests that our architecture not only preserves its ability to detect seizures accurately but also maintains a balanced trade-off between precision and recall. It is crucial for real-world applications where false positives and negatives have significant implications. These results underscore the resilience of the proposed model to structural modifications, highlighting its adaptability and efficiency in resource-constrained environments. The ability of the model to maintain high-performance levels even with reduced computational complexity holds promising implications for practical deployment.

### C. Out-of-sample Continental Generalization

*Can the KAN-EEG generalize beyond in-sample testing?* To evaluate generalization performance, we tested our initial model with 2 layers (764 and 256 neurons respectively) on two distinct out-of-sample datasets. The RPAH dataset results are presented in Table III, and the EPILEPSIAE dataset results are shown in Table IV. The results shown demonstrate some out-of-sample testing ability, albeit not efficiently in both the RPAH and EPILEPSIAE dataset with AUROC results of 0.60 and 0.55 respectively.

**TABLE III.**
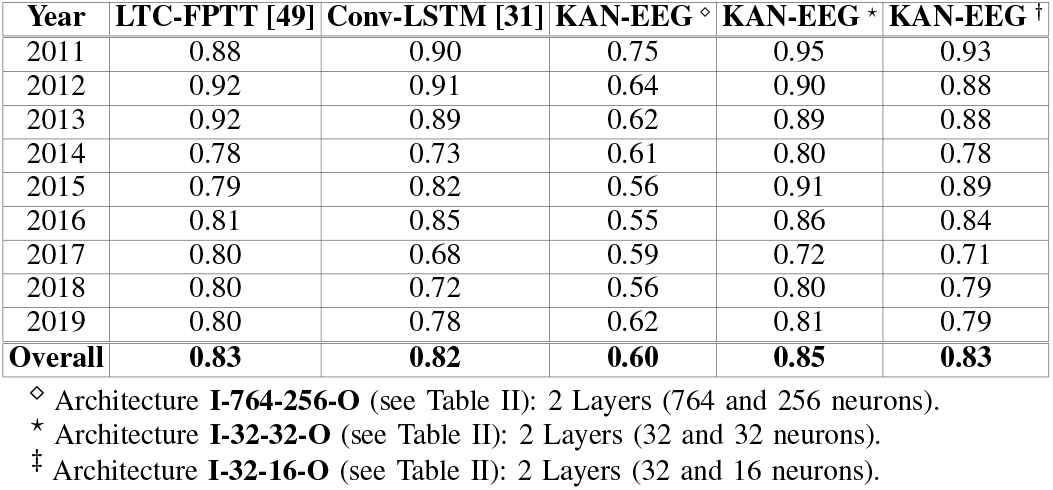
Out-of-sample generalization results (AUROC) on the RPAH dataset.

**TABLE IV.** Out-of-sample generalization results (AUROC) on the EPILEPSIAE dataset.

| Pat-ID | LTC-FPTT [49] | Conv-LSTM [31] | KAN-EEG <sup>◊</sup> | KAN-EEG <sup>*</sup> | KAN-EEG <sup>†</sup> |
| --- | --- | --- | --- | --- | --- |
| 1 | 0.88 | 0.88 | 0.63 | 0.84 | 0.79 |
| 2 | 0.82 | 0.88 | 0.49 | 0.58 | 0.55 |
| 3 | 0.84 | 0.85 | 0.60 | 0.85 | 0.77 |
| 4 | 0.82 | 0.89 | 0.31 | 0.71 | 0.58 |
| 5 | 0.79 | 0.72 | 0.58 | 0.73 | 0.72 |
| 6 | 0.78 | 0.69 | 0.42 | 0.64 | 0.60 |
| 7 | 0.78 | 0.81 | 0.53 | 0.75 | 0.76 |
| 8 | 0.79 | 0.81 | 0.63 | 0.88 | 0.82 |
| 9 | 0.81 | 0.92 | 0.65 | 0.90 | 0.88 |
| 10 | 0.81 | 0.68 | 0.43 | 0.57 | 0.49 |
| 11 | 0.81 | 0.81 | 0.82 | 0.96 | 0.95 |
| 12 | 0.81 | 0.74 | 0.48 | 0.75 | 0.59 |
| 13 | 0.83 | 0.86 | 0.33 | 0.78 | 0.63 |
| 14 | 0.83 | 0.85 | 0.65 | 0.84 | 0.82 |
| 15 | 0.83 | 0.71 | 0.58 | 0.81 | 0.82 |
| 16 | 0.83 | 0.85 | 0.77 | 0.93 | 0.91 |
| 17 | 0.83 | 0.83 | 0.44 | 0.65 | 0.63 |
| 18 | 0.83 | 0.82 | 0.52 | 0.82 | 0.68 |
| 19 | 0.84 | 0.91 | 0.74 | 0.88 | 0.87 |
| 20 | 0.84 | 0.68 | 0.34 | 0.71 | 0.65 |
| 21 | 0.84 | 0.83 | 0.47 | 0.83 | 0.71 |
| 22 | 0.84 | 0.95 | 0.62 | 0.74 | 0.70 |
| 23 | 0.85 | 0.84 | 0.60 | 0.89 | 0.84 |
| 24 | 0.84 | 0.83 | 0.76 | 0.85 | 0.82 |
| 25 | 0.84 | 0.62 | 0.56 | 0.75 | 0.73 |
| 26 | 0.84 | 0.84 | 0.38 | 0.67 | 0.57 |
| 27 | 0.84 | 0.86 | 0.60 | 0.71 | 0.70 |
| 28 | 0.84 | 0.91 | 0.70 | 0.88 | 0.88 |
| 29 | 0.84 | 0.72 | 0.51 | 0.79 | 0.76 |
| 30 | 0.85 | 0.88 | 0.43 | 0.75 | 0.62 |
| Overall | **0.83** | **0.81** | **0.55** | **0.78** | **0.73** |

<sup>◊</sup> Architecture **I-764-256-O** (see Table II): 2 Layers (764 and 256 neurons).
<sup>\*</sup> Architecture **I-32-32-O** (see Table II): 2 Layers (32 and 32 neurons).
<sup>†</sup> Architecture **I-32-16-O** (see Table II): 2 Layers (32 and 16 neurons).

The observed results primarily stem from an architectural design showing indications of overfitting on the training dataset. This assertion suggests that the model has become overly specialized in capturing specific information inherent in the training/validation data, consequently compromising its ability to generalize in different datasets. As a result, addressing overfitting is crucial for enhancing the model’s robustness and applicability in real-world scenarios, with unseen-world datasets with different seizure patterns that can be specific to each patient.

### D. Promises for compact networks lead to better performance

To address the challenges of inefficient generalization, we modified the model to have two layers with 32 and 16 neurons, respectively. Additionally, we provided an extra evaluation to confirm that our findings are assertive by testing in a model with two layers, each with 32 neurons. As these architectures showed robustness in in-sample testing, we decided to apply them to the generalization test using weights from training when the AUROC metric was not increasing.

As discussed previously, the KAN-EEG model with two layers (764 and 256 neurons) had a lower AUROC than the LTC-FPTT and ConvLSTM models. Interestingly, the KAN-EEG models with two layers (32 and 16 neurons, and 32 and 32 neurons) demonstrated comparable or superior AUROC values relative to these models. This suggests that overfitting may occur in models with more complex neuron structures, adversely affecting their generalization performance.

These findings are significant as they indicate the potential for future deployable on-edge KAN algorithms, where memory size compression and robustness are crucial. These findings suggest the potential for replacing MLP-based models for seizure detection with the KAN-EEG algorithm. Our framework remains robust and demonstrates promise for practical implementation in real-world applications. It is important to highlight that our data was not trained on the full TUH dataset, indicating that there is even further room for improvement in achieving greater results.

## IV. Discussion

In this study, we pioneer the application of shallow KANs to EEG data, leveraging this model’s unique structure and capabilities inspired by the Kolmogorov-Arnold representation theorem. The proposed architecture not only achieves a higher accuracy in-sample dataset but can also generalized across different scalp-EEG, as smaller KANs can outperform much larger MLPs in terms of data fitting and partial differential equation solving, owing to faster neural scaling laws [12]. By applying KANs to EEG data, we expect to unlock new potentials in seizure prediction models, given its ability to learn compositional structures and optimize univariate functions [12]. This novel application could provide more accurate and interpretable predictions, advancing the neural network-based seizure prediction field. The model’s resilience against catastrophic forgetting will also ensure stable and continuous learning, a critical requirement for medical applications involving long-term EEG monitoring. To this extent, our next steps will involve the deployment of this architecture on memristor devices.

## V. Conclusion

This study has introduced the KANs as a novel approach for epileptic seizure detection using EEG signals. By leveraging its unique architecture and learning properties, which feature learnable activation functions on edges instead of fixed ones on nodes, we have demonstrated the potential for achieving comparable accuracy and higher accuracy in both in-sample and out-of-sample seizure detection compared to traditional MLP models. Our results indicate that KANs should be a solution for replacing MLPs-based models for seizure detection applications, requiring a smaller network size while maintaining robust performance. The significant advancements presented in this work highlight the potential of KANs to improve clinical outcomes for patients with epilepsy by providing a more accurate, efficient, and interpretable tool for seizure detection that enhances decision-making, treatment planning, and overall disease management.

## Data Availability

The TUH dataset is publicly available https://isip.piconepress.com/projects/tuh_eeg/html/downloads.shtml. The EPILEPSIAE dataset is available at cost via in http://www.epilepsiae.eu/project_outputs/european_database_on_epilepsy. The Department of Neurology at the Royal Prince Alfred Hospital (RPAH) dataset was used under Ethics Review Board approval (NSW (New South Wales), Local Health District (LHD) Ethics X19-0323-2019/STE16040) and is not publicly available

http://www.epilepsiae.eu/project_outputs/european_database_on_epilepsy

## VI. Acknowledgement

Luis Fernando Herbozo Contreras would like to acknowledge the partial support of the Faculty of Engineering Research Scholarship provided by the University of Sydney. Zhaojing Huang would like to acknowledge the support of the Research Training Program (RTP) provided by the Australian Government. Omid Kavehei acknowledges the support provided by The University of Sydney through a SOAR Fellowship and Microsoft’s support through a Microsoft AI for Accessibility grant.

## VII. Ethics declaration

No ethics declaration was needed for the TUH and EPILEPSIAE dataset. For this study, NSW Local Health District (LHD) ethics X19-0323-2019/STE16040 is approved for using the RPAH dataset in collaboration between The University of Sydney and the Comprehensive Epilepsy Services, the Department of Neurology, at the Royal Prince Alfred Hospital, Australia.

## VIII. Data Accessibility

The TUH dataset is publicly available here. The EPILEPSIAE dataset is available at cost via this link. The Department of Neurology at the Royal Prince Alfred Hospital (RPAH) dataset was used under Ethics Review Board approval and is not publicly available.

